# Intention to receive a COVID-19 vaccine: Results from a population-based survey in Canada

**DOI:** 10.1101/2021.02.03.21251007

**Authors:** Gina S. Ogilvie, Shanlea Gordon, Laurie W. Smith, Arianne Albert, C. Sarai Racey, Amy Booth, Anna Gottschlich, David Goldfarb, Melanie C.M. Murray, Liisa A.M. Galea, Angela Kaida, Lori A. Brotto, Manish Sadarangani

**Affiliations:** BC Centre for Disease Control, Provincial Health Services Authority, 655 West 12^th^ Avenue, Vancouver, British Columbia, V5Z 4R4; Faculty of Medicine, University of British Columbia, 2329 West Mall, Vancouver, British Columbia, Canada, V6T 1Z4; Women’s Health Research Institute, 4500 Oak Street, Vancouver, British Columbia, Canada, V6H 2N9; BC Children’s Hospital Research Institute, 938 W 28^th^ Avenue, Vancouver, British Columbia, Canada, V5Z 4H4; BC Cancer Agency, 600 W 10^th^ Avenue, Vancouver, British Columbia, Canada, V5Z 4E6; Department of Pathology and Laboratory Medicine, BC Children’s and Women’s Health Centre, 4500 Oak Street, Vancouver, British Columbia, Canada, V6H 3N1; Faculty of Arts, University of British Columbia, 2329 West Mall, Vancouver, British Columbia, Canada, V6T 1Z4; Faculty of Health Sciences, Simon Fraser University, 8888 University Drive, Burnaby, British Columbia, V5A 1S6; Vaccine Evaluation Centre, 950 W 28^th^ Avenue, Vancouver, British Columbia, Canada, V5Z 4H4

**Keywords:** COVID-19, Vaccine hesitancy, Vaccine confidence, Vaccine, Canada, Public health

## Abstract

**Background:** The success of any COVID-19 vaccine program ultimately depends on high vaccine uptake. This study determined overall intention to receive a COVID-19 vaccine and identified factors that predict intentions to be vaccinated against COVID-19 in Canada, specifically in key priority groups identified by the American Committee on Immunization Practice (ACIP) and the National Advisory Committee on Immunization (NACI) for early immunization.

**Methods:** Individuals from research cohorts from the general population of British Columbia aged 25-69 were invited complete an online survey based on validated scales and theoretical frameworks to explore intention to receive a COVID-19 vaccine. Two multivariable logistic regression models were conducted to determine factors associated with intention to receive the COVID-19 vaccine.

**Results:** Of 4,948 respondents, 79.8% intended to receive a COVID-19 vaccine. In multivariable modeling, respondents who intended to receive the vaccine had higher vaccine attitudinal scores (p <0.001), reported greater influence of direct social norms (p = 0.001), and indirect social norms, including their family physician (p = 0.024), and Provincial Health Officer (p = 0.011). Older individuals (>60 years) were more likely to intend to receive the vaccine, while females (95%CI 0.57,0.93), those with less than high school education (95%CI 0.5,0.76), those who self-identified as non-white (95%CI 0.60,0.92), self-identified as Indigenous (95%CI 0.36,0.84) and essential non-health care workers (95%CI 0.59,0.86) had lower adjusted odds of intending to receive a COVID-19 vaccine.

**Conclusions:** To optimize vaccine coverage, public health should focus on key messages around vaccine safety and benefit, and leverage trusted practitioners for messaging. As certain key populations identified by NACI and ACIP for early immunization report a lower intention to vaccinate, there is a need for in-depth education and support for these communities to ensure optimal uptake.

## Background

The development of safe and effective COVID-19 vaccines is a critical step in ending the pandemic.^1,2^ Across Canada vaccine distribution has commenced. The Pfizer and Moderna vaccines, both of which show approximately 95% protection against COVID-19^3,4^, are currently being distributed in each province, including British Columbia. Global health authorities including the World Health Organization (WHO)^5^ and the American Committee on Immunization Practice (ACIP)^6^ have provided guidance for vaccine roll-out globally with the acknowledgment that initial vaccine supply will be limited. In Canada, the National Advisory Committee on Immunization (NACI) has identified priority populations for initial vaccine roll-out, including populations at high risk for severe COVID-19 related illness; those most likely to transmit COVID-19 to those at high risk; those essential to maintaining the COVID-19 response; those who contribute to the maintenance of essential services; and those living or working in conditions that put them at higher risk for infection.^7^ ACIP had identified similar priority populations for early vaccine distribution in the United States, including healthcare personnel, persons with high-risk underlying medical conditions, and individuals over 65 years^6^.

Ultimately, the success of any COVID-19 vaccine program depends upon vaccine uptake in the population. Over the past decade there has been a significant rise in “vaccine hesitancy”, a complex concept defined as the refusal, reluctance, or delay in acceptance to vaccinate despite vaccine availability^2,8,9^, which has led to decreases in vaccine uptake.^10,11,12,13^ Understanding the predictors and determinants influencing intentions to receive a COVID-19 vaccine in Canada are key to designing public health programming to optimize vaccination rates, including among priority populations, when vaccine is available broadly.^6,7^ When evaluating vaccine intention, assessments should be based on validated theoretical frameworks in order to provide robust information for program planning. Using a survey based on the WHO Vaccine Hesitancy Scale (VHS) and grounded in the Theory of Planned Behaviour (TPB), the primary objective of this study was to determine the intention to receive a COVID-19 vaccine among people living in British Columbia. The secondary objectives were to identify factors that predict intentions to be vaccinated, specifically in priority groups identified by NACI, in order to guide public health vaccination programs.

British Columbia is the western most province in Canada, and has a population of more than 5 million. Over 80% of the population lives in a population centre^14^. Building on a well-established vaccine program, COVID-19 vaccine distribution in BC will be centrally organized by the BC Center for Disease Control, with regional delivery coordinated by regional Health Authorities^15^.

## Methods

The current study is part of a larger, ongoing investigation led by the Women’s Health Research Institute evaluating the impacts of COVID-19 and public health controls on British Columbians.^16^

### Participants and recruitment

Prospective participants were part of large research cohorts from the general population of British Columbia (BC) who had consented to be contacted for future research. Eligible individuals (aged 25-69; resident of BC) were sent an email invitation to participate (*Index Participants*) in an online survey. To increase diverse sex and gender representation, respondents were asked to provide the email address of an adult household member who identified as another gender to participate in the survey – these individuals were then invited to participate in the online survey (*Household Participants*). All prospective participants received up to three email reminders, and an opportunity to participate in a draw for a gift card. Participants could also opt-in to receive an at-home SARS-CoV-2 (severe acute respiratory syndrome coronavirus 2) research antibody test (data collection ongoing and will be reported separately). Ethical approval was received from The University of British Columbia Research Ethics Board (H20-01421). All methods performed as a part of this study were in accordance with the UBC Research Ethics Board guidelines. Informed consent was obtained from all participants prior to participation in this study.

### Survey Design and Measures

To determine the factors associated with intention to be vaccinated, existing items came from the validated WHO VHS^9^ and new items were developed using TPB framework.^17^ TPB is a psychological model of behavior change that has been used widely to predict and understand health-related behaviours and has been shown previously to accurately predict vaccine uptake in Canada.^18,19,20,21,22^ TPB defines the most significant predictors of a health behaviour as attitudes, social norms (direct, indirect), and perceived behavioural controls.^17^ For this survey, items were developed from a detailed literature review and through elicitation surveys of experts to identify key factors expected to influence intention to receive a COVID-19 vaccine (Additional File 1). The survey assessed vaccine attitudes (8 items), direct social norms (4 items), indirect social norms (14 items), and perceived behavioural controls (4 items) (Additional File 2). Vaccine hesitancy was measured using the validated 9-item, 2-factor VHS initially developed by the WHO Sage Working Group on Vaccine Hesitancy8,9,12, and adapted for BC. The factors included in VHS are vaccine lack of confidence (7 items) and vaccine risk (2 items).^8^ All items in TPB and VHS scales were measured on a 5-point Likert scale. The overall survey was assessed for face validity and comprehension, pilot tested, and the final version was implemented using REDCap (Research Electronic Data Capture).^23^

For each participant the following demographic characteristics were assessed: age, sex, gender, Indigenous ancestry, visible minority status, education, household composition, existing chronic health condition, self-reported history of COVID-19, and self-reported employment as an essential worker. Visible minority categories were based on the Statistics Canada 2016 census. The primary outcome was response to “*If a COVID-19 vaccine were to become available to the public, and recommended for you, how likely are you to receive it*?”

### Survey response rate

Response rate was calculated according to the American Association for Public Opinion Research (AAPOR) guidelines for Internet Surveys of Specifically Named Persons.^24^ Response rate is defined as the sum of complete (100% of applicable questions answered) and partial surveys (<100% of applicable questions answered), divided by the overall number of invitations distributed to eligible respondents. Participant disposition is defined as: respondents (number of complete and partial surveys), ineligible (those who did not meet the eligibility criteria), invitation returned undelivered (number of emails bounced back), explicit refusals (those who replied that they did not want to participate), implicit refusals (those who visited the online survey but failed to complete any survey items), and nothing ever returned (those who did not respond to the survey invites).^24^ With a sample size of 4,500, we had 80% power to detect +/-1.34% 95%CI around an estimated overall vaccine acceptance rate of 70%.^25^ We plan to compare ages of responders and non-responders to determine representativeness of the study participants.

### Analyses

Analyses were carried out in R v.4.0.2.^26^ Mean values for TPB and VHS scales were calculated. Item reliability for TPB scales was assessed using Cronbach’s α, and if α >0.6 (good agreement), scales were included in bivariable and multivariable analysis. For the primary outcome (intention to receive a COVID-19 vaccine) responses on the 5-point Likert scale were dichotomized, with those who responded *very* or *somewhat likely* coded as “intending to vaccinate” and those who responses *neutral, unlikely*, or *very unlikely* coded as “not intending to vaccinate”. We investigated the relationship between intention to get a COVID-19 vaccine and demographic and vaccine specific variables using mixed-effects logistic regressions. To allow participants to select more than one visible minority category, visible minority variables were coded as those who indicated a particular minority status vs. those who did not, and used as separate variables in analyses. Intention to vaccinate was examined in priority groups identified by ACIP and NACI, including older (>60), those with chronic health conditions, visible minorities, Indigenous participants, healthcare workers, and non-healthcare essential workers.^7^ Two multivariable mixed-effects logistic regression models were conducted to explore factors associated with the dependent variable ‘*intention to receive a COVID-19 vaccine’*. One model examined demographic variables, and the other model included the VHS and TPB items. For both models, *a priori* predictors of vaccine intention which achieved a p<0.1 from bivariable analysis were included in multivariable models, and adjusted odds ratios were calculated to identify factors associated with intention to receive the COVID-19 vaccine. Case-wise deletion was used to address missing data within each bivariable analysis. Multivariable analyses included only non-missing data. Clustering by household was accounted for in the mixed-effects models using a random effect for household identification. A descriptive analysis of male versus female respondents was completed for the WHO VHS scale and the TPB framework.

## RESULTS

### Survey Respondents

Between August 20 and September 27, 2020, 13,764 survey invites were distributed to prospective *Index Participants* and 4,292 responded to the survey (Figure 1). Of the 1291 invites that were forwarded to prospective *Household Participants*, 656 responded. Overall, 4,058 surveys were completed and 890 were partially completed by eligible participants, for a response rate of 32.9% (4,948/15,055) overall (including all who were *sent* the survey), and 37.2% (4,948/13,299) (including only those who *received* the survey). We compared the ages of survey respondents and non-respondents (those who declined participation or did not respond) across five age strata (25-29; 30-39; 40-49; 50-59; 60-69). We found no significant differences between the ages of respondents and non-respondents, indicating representativeness of our sample based on age (data not shown).

**Figure 1:**
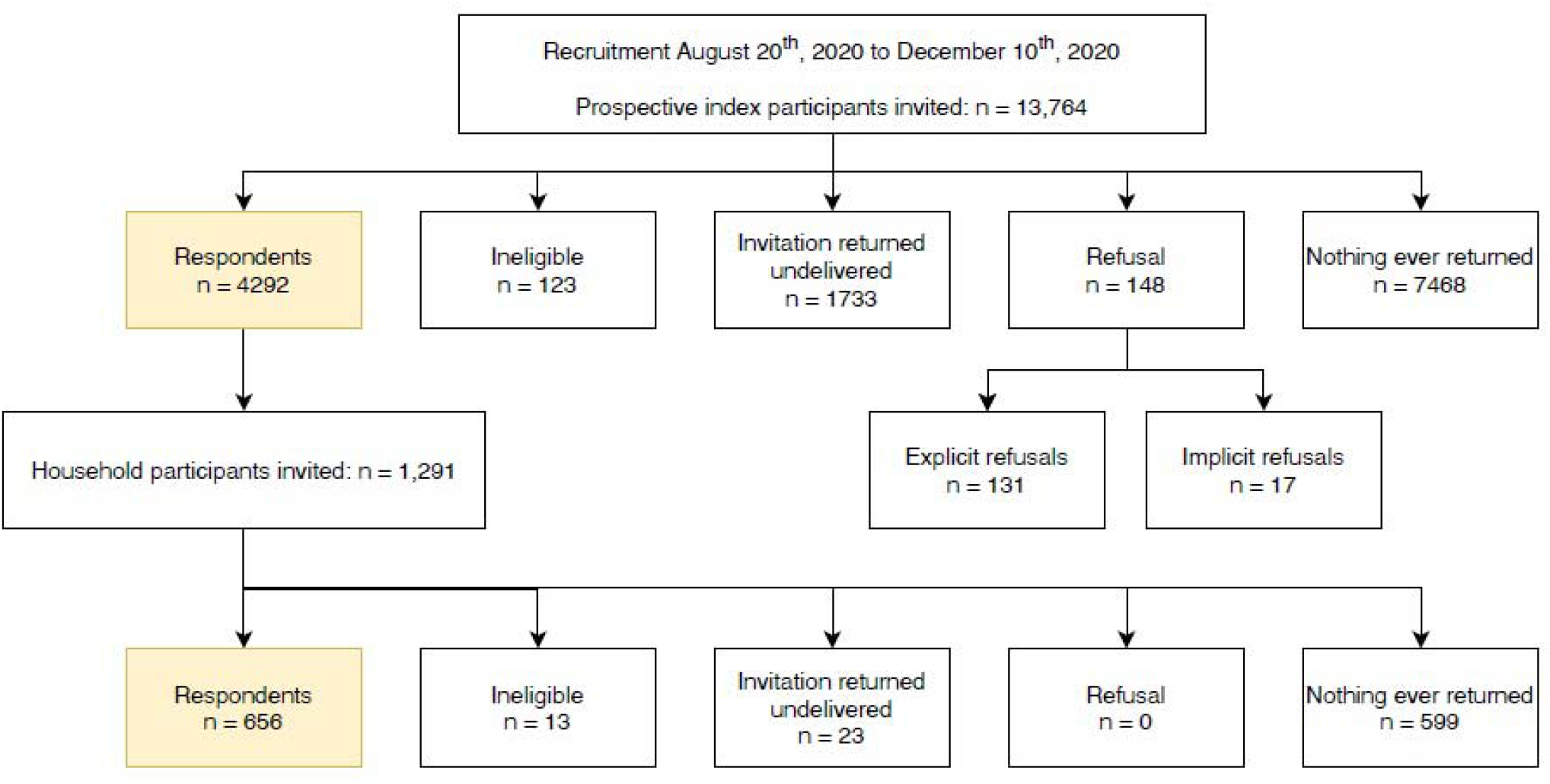
Study Participant CONSORT Diagram^1^. ^1^**American Association for Public Opinion Research:** *Respondents:* number of complete and partial surveys *Ineligible:* those who did not meet the eligibility criteria (aged 25-69; resident of BC) *Invitation returned undelivered*: number of emails bounced back *Refusal:* those who declined to participate *Explicit refusals*: those who replied that they did not want to participate *Implicit refusals*: those who visited the survey URL but failed to complete any survey items *Nothing ever returned*: those who did not respond to the survey invites

### Demographics

Survey respondents had a mean age of 51.8 (SD= 10.5) with a range from 25-69 years (Table 1). The majority of participants self-reported being assigned female sex at birth (84.8%), identified as women (84.1%), white (82.6%), and had more than a high school education (83.8%). Fifty-two participants (1.1%) identified as non-binary, GenderQueer, Agender, Two-Spirit, or other gender identity. Visible minority responses were grouped into broader categories for analyses. Essential health care workers comprised 12.0% of respondents and essential non-healthcare workers comprised 18.4%. Survey respondents were from all five health authorities in BC.

**Table 1:** Respondent Demographics (N=4,948)

|  | **Total**<br>**N = 4948** |
| --- | --- |
| **Age** |  |
| 25-29 | 111 (2.2%) |
| 30-39 | 573 (11.6%) |
| 40-49 | 1260 (25.5%) |
| 50-59 | 1496 (30.2%) |
| 60-69 | 1370 (27.7%) |
| Missing | 138 (2.8%) |
| **Sex** |  |
| Female | 4196 (84.8%) |
| Male | 609 (12.3%) |
| Missing | 143 (2.9%) |
| **Gender** |  |
| Woman | 4159 (84.1%) |
| Man | 597 (12.1%) |
| Non-Binary, GenderQueer, Agender, Two-spirit,<br>or other | 52 (1.1%) |
| Missing | 140 (2.8%) |
| **Indigenous** |  |
| Indigenous | 127 (2.6%) |
| Not Indigenous | 4467 (90.3%) |
| Prefer not to answer | 34 (0.7%) |
| Missing | 320 (6.5%) |
| **Asian<sup>1</sup>** |  |
| No | 4563 (92.2%) |
| Yes | 360 (7.3%) |
| Missing | 25 (0.5%) |
| **Latin American** |  |
| Latin American | 65 (1.3%) |
| Not | 4858 (98.2%) |
| Missing | 25 (0.5%) |
| **South Asian<sup>2</sup>** |  |
| No | 4826 (97.5%) |
| Yes | 97 (2.0%) |
| Missing | 25 (0.5%) |
| **Black** |  |
| No | 4895 (98.9%) |
| Yes | 28 (0.6%) |
| Missing | 25 (0.5%) |
| **White** |  |
| Yes | 4089 (82.6%) |
| No | 834 (16.9%) |
| Missing | 25 (0.5%) |
| **Education** |  |
| More than High School | 4148 (83.8%) |
| High School or less | 655 (13.2%) |
| Missing | 145 (2.9%) |
| **Essential worker** |  |
| No | 3302 (66.7%) |
| Yes, health worker | 594 (12.0%) |
| Yes, other essential worker | 910 (18.4%) |
| Missing | 142 (2.9%) |
| **Number of Adults in Household** |  |
| One | 1151 (23.3%) |
| Two | 2598 (52.5%) |
| Three or more | 1,050 (21.2%) |
| Missing | 149 (3.0%) |
| **Children < 5** |  |
| None | 4349 (87.9%) |
| One | 273 (5.5%) |
| Two or more | 83 (1.7%) |
| Missing | 243 (4.9%) |
| **Children 5-17** |  |
| None | 3242 (65.5%) |
| One | 731 (14.8%) |
| Two or more | 760 (15.4%) |
| Missing | 215 (4.3%) |
| **Chronic Health Conditions** |  |
| None | 2302 (46.5%) |
| One or more | 2490 (50.3%) |
| Missing | 156 (3.2%) |
| **Do you think you had COVID-19** |  |
| No | 4290 (86.7%) |
| Yes | 534 (10.8%) |
| Missing | 124 (2.5%) |
| **WHO scale: Lack of Confidence in Vaccines** |  |
| Mean (SD) | 1.3 ($\pm 0.6$) |
| Missing | 170 (3.4%) |
| **WHO scale: Vaccine Risks** |  |
| Mean (SD) | 3.0 ($\pm 1.1$) |
| Missing | 173 (3.5%) |
| **Attitudes to COVID-19 Vaccine** |  |
| Mean (SD) | 34.6 ($\pm 5.8$) |
| Missing | 609 (12.3%) |
| **Perceived Behavioural Control** |  |
| Mean (SD) | 16.0 ($\pm 2.6$) |
| Missing | 509 (10.3%) |
| **Direct Social Norms** |  |
| Mean (SD) | 14.8 ($\pm 3.3$) |
| Missing | 586 (11.8%) |
| **Indirect Social Norms: Family Doctor/PHCP** |  |
| Mean (SD) | 6.0 ($\pm 3.6$) |
| Missing | 563 (11.4%) |
| **Indirect Social Norms: BC Provincial Health Officer** |  |
| Mean (SD) | 6.7 ($\pm 3.6$) |
| Missing | 562 (11.4%) |
| **Indirect Social Norms: Coworkers** |  |
| Mean (SD) | 4.4 ($\pm 3.5$) |
| Missing | 1797 (36.3%) |
| **Indirect Social Norms: Employer** |  |
| Mean (SD) | 5.2 ($\pm 3.4$) |
| Missing | 1777 (35.9%) |
| **Indirect Social Norms: Educational Institution** |  |
| Mean (SD) | 4.7 ($\pm 3.4$) |
| Missing | 2899 (58.6%) |
| **Indirect Social Norms: Friends** |  |
| Mean (SD) | 4.1 ($\pm 3.3$) |
| Missing | 556 (11.2%) |
| **Indirect Social Norms: Family** |  |
| Mean (SD) | 5.5 ($\pm 3.7$) |
| Missing | 528 (10.7%) |
| **Indirect Social Norms: Total** |  |
| Mean (SD) | 22.4 ($\pm 11.8$) |
| Missing | 672 (13.6%) |

<sup>1</sup> Asian: Chinese, Korean, Japanese, Filipino, Laotian, Cambodian, Vietnamese, etc.
<sup>2</sup> South Asian: Indian, Pakistani, Bangladeshi, Sri Lankan, Bhutanese, Nepalese, etc.

### Intention to receive a COVID-19 vaccine

Overall, 79.8% were ‘*somewhat* or *very likely’* to receive a COVID-19 vaccine if it was available to the public and recommended for them, adjusted for household clustering (Table 2). Among essential health care workers, 81.8% indicated that they intend to receive a COVID-19 vaccine. In bivariable analyses, those who were older (>60 years), males, had chronic health conditions, were essential health care workers, had more than a high school education, or had two adults in the house were significantly more likely to intend to receive the COVID-19 vaccine (p≤0.05). Specifically, participants in all age groups except 25-29 were significantly less likely to intend to receive the COVID-19 vaccine compared to those in the 60-69 group. Individuals who were essential non-health care workers, identified as non-white, South Asian or of Indigenous ancestry were significantly less likely to intend to receive vaccination (p≤0.05). Given there were fewer than 52 non-binary, GenderQueer, Agender, or Two-spirit respondents, sex, not gender, was used in the model. In multivariable modeling, individuals who were older (>60 years) were more likely to intend to receive the COVID-19 vaccine (Table 2), while females (AOR 0.7; 95%CI 0.55, 0.89); those who had less than a high school education (AOR 0.62; 95%CI 0.51, 0.77); those who self-identified as non-white (AOR 0.76; 95%CI 0.61, 0.95); those who self-identified as Indigenous (AOR 0.58; 95%CI 0.38, 0.87); those who were essential non-health care workers (AOR 0.72; 95%CI 0.6, 0.87); and those who thought they had COVID-19 (AOR 0.76; 95% 0.61 – 0.96) had lower adjusted odds of intending to receive a COVID-19 vaccine if it were publicly available and recommended for them.

**Table 2:** Bivariable comparison and multivariable model for demographic factors associated with intention to receive COVID-19 vaccine (N=4787)

|  | Likely to receive COVID-19 vaccine –<br>Bivariable comparisons* |  |  |  | Likely to receive COVID-19 vaccine -<br>Multivariable model |  |  |
| --- | --- | --- | --- | --- | --- | --- | --- |
|  | % (95%CI)* | OR | CI | P-value** | OR | CI | P-value** |
| **Age** |  |  |  |  |  |  |  |
| 60-69 | 84.2 (82.1 - 86.1) | *reference* |  |  | *reference* |  |  |
| 25-29 | 85.6 (77.6 - 91.0) | 1.12 | 0.64 - 1.95 | 0.7 | 1.42 | 0.77 - 2.64 | 0.26 |
| 30-39 | 75.5 (71.7 - 79.0) | 0.58 | 0.45 - 0.74 | <0.0001 | 0.64 | 0.49 - 0.83 | 0.0008 |
| 40-49 | 78.8 (76.4 - 81.1) | 0.7 | 0.57 - 0.86 | 0.0006 | 0.78 | 0.62 - 0.97 | 0.02 |
| 50-59 | 77.8 (75.6 - 79.9) | 0.66 | 0.54 - 0.80 | <0.0001 | 0.67 | 0.55 - 0.82 | 0.0001 |
| **Sex** |  |  |  |  |  |  |  |
| Male | 84.6 (81.4 - 87.3) | *reference* |  |  | *reference* |  |  |
| Female | 79.2 (77.9 - 80.4) | 0.7 | 0.55 - 0.88 | 0.003 | 0.7 | 0.55 - 0.89 | 0.004 |
| Missing |  |  |  |  |  |  |  |
| **Gender** |  |  |  |  |  |  |  |
| Woman | 79.0 (77.7 - 80.3) | *reference* |  |  |  |  |  |
| Man | 84.7 (81.5 - 87.4) | 1.47 | 1.16 - 1.87 | 0.002 |  |  |  |
| Non-Binary, GenderQueer,<br>Agender, Two-spirit, or other | 92.0 (80.3 - 97.0) | 3.04 | 1.08 - 8.55 | 0.04 |  |  |  |
| **Indigenous** |  |  |  |  |  |  |  |
| Not Indigenous | 80.5 (79.2 - 81.6) | *reference* |  |  | *reference* |  |  |
| Indigenous | 66.5 (57.5 - 74.4) | 0.48 | 0.32 - 0.71 | 0.0002 | 0.58 | 0.38 - 0.87 | 0.009 |
| **Asian** |  |  |  |  |  |  |  |
| No | 80.1 (78.8 - 81.3) | *reference* |  |  |  |  |  |
| Yes | 76.7 (71.9 - 80.9) | 0.82 | 0.63 - 1.07 | 0.14 |  |  |  |
| **South Asian** |  |  |  |  |  |  |  |
| No | 80.1 (78.9 - 81.2) | *reference* |  |  | *reference* |  |  |
| Yes | 66.5 (56.2 - 75.5) | 0.49 | 0.32 - 0.77 | 0.002 | 0.65 | 0.39 - 1.07 | 0.09 |
| **Latin American** |  |  |  |  |  |  |  |
| No | 79.9 (78.7 - 81.0) | *reference* |  |  |  |  |  |
| Yes | 75.3 (63.1 - 84.5) | 0.77 | 0.43 - 1.37 | 0.37 |  |  |  |
| **Black** |  |  |  |  |  |  |  |
| No | 79.9 (78.7 - 81.0) | reference |  |  |  |  |  |
| Yes | 67.7 (48.1 - 82.6) | 0.53 | 0.23 - 1.20 | 0.13 |  |  |  |
| **White** |  |  |  |  |  |  |  |
| Yes | 80.8 (79.6 - 82.0) | reference |  |  | reference |  |  |
| No | 73.8 (70.3 - 77.0) | 0.67 | 0.55 - 0.81 | < 0.0001 | 0.76 | 0.61 - 0.95 | 0.01 |
| **Education** |  |  |  |  |  |  |  |
| More than High School | 80.8 (79.6 - 82.0) | reference |  |  | reference |  |  |
| High School or less | 73.2 (69.6 - 76.6) | 0.65 | 0.53 - 0.79 | < 0.0001 | 0.62 | 0.51 - 0.77 | <0.0001 |
| **Essential worker** |  |  |  |  |  |  |  |
| No | 80.9 (79.5 - 82.3) | reference |  |  | reference |  |  |
| Yes, health worker | 81.8 (78.5 - 84.8) | 1.06 | 0.84 - 1.34 | 0.61 | 1.1 | 0.86 - 1.40 | 0.45 |
| Yes, other essential worker | 74.5 (71.5 - 77.3) | 0.69 | 0.58 - 0.82 | <0.0001 | 0.72 | 0.60 - 0.87 | 0.001 |
| **Chronic Health Conditions** |  |  |  |  |  |  |  |
| No chronic conditions | 78.6 (76.9 - 80.3) | reference |  |  | reference |  |  |
| Any chronic condition | 80.9 (79.3 - 82.5) | 1.15 | 1.00 - 1.33 | 0.05 | 1.14 | 0.98 - 1.32 | 0.10 |
| **Number of Adults in Household** |  |  |  |  |  |  |  |
| One | 78.4 (75.9 - 80.8) | reference |  |  |  |  |  |
| Two | 81.3 (79.7 - 82.8) | 1.2 | 1.00 - 1.43 | 0.05 |  |  |  |
| Three or more | 77.8 (75.1 - 80.3) | 0.96 | 0.78 - 1.19 | 0.72 |  |  |  |
| **Children < 5** |  |  |  |  |  |  |  |
| None | 80.1 (78.9 - 81.3) | reference |  |  |  |  |  |
| One | 77.2 (66.7 - 85.1) | 0.87 | 0.64 - 1.18 | 0.36 |  |  |  |
| Two or more | 77.8 (72.3 - 82.4) | 0.84 | 0.49 - 1.43 | 0.52 |  |  |  |
| **Children 5-17** |  |  |  |  |  |  |  |
| None | 80.3 (78.9 - 81.7) | reference |  |  |  |  |  |
| One | 78.6 (75.4 - 81.5) | 0.9 | 0.74 - 1.11 | 0.32 |  |  |  |
| Two or more | 79.6 (76.5 - 82.4) | 0.96 | 0.78 - 1.17 | 0.67 |  |  |  |
| **Do you think you had COVID-19** |  |  |  |  |  |  |  |
| No | 80.3 (79.0 - 81.5) | reference |  |  | reference |  |  |
| Yes | 76.2 (72.3 - 79.7) | 0.79 | 0.63 - 0.98 | 0.03 | 0.76 | 0.61 - 0.96 | 0.02 |

\* Estimated conditional proportion: effects models
\*\* P-values from Wald tests against reference category

### Vaccine Hesitancy Scale and Theory of Planned Behaviour

All items in the TPB scale had good to strong agreement (Cronbach’s alpha>0.6) and were included in the analysis (Table 3). In bivariable modeling, all responses in the WHO and TPB scales were significantly different in individuals who intended to be vaccinated and those who did not, and were included in the multivariable analysis (Table 4). In multivariable modeling (Table 4), participants who had higher vaccine confidence and who perceived a lower vaccine risk were more likely to intend to receive a COVID-19 vaccine (p < 0.001). In the TPB scales, respondents who intended to vaccinate had significantly higher vaccine attitudinal scores (p <0.001), reported greater influence of direct social norms on their decision to vaccinate, including belief that most people who are important to them would think they should receive the COVID-19 vaccine, and would expect them to receive the vaccine (p < 0.001). Participants intending to be vaccinated were also significantly more likely to report greater influence of indirect social norms, including the opinions of family (p < 0.001), their family physician or primary healthcare provider (p = 0.03), and the Provincial Health Officer (p = 0.01). Perceived behavioural controls and the influence of friends (indirect social norm) were not found to be predictors of intention to vaccinate. A descriptive analysis of male (n = 605) versus female (n = 4,178) respondents was completed for the TPB scale and the WHO VHS scale (Table 5).

**Table 3:**
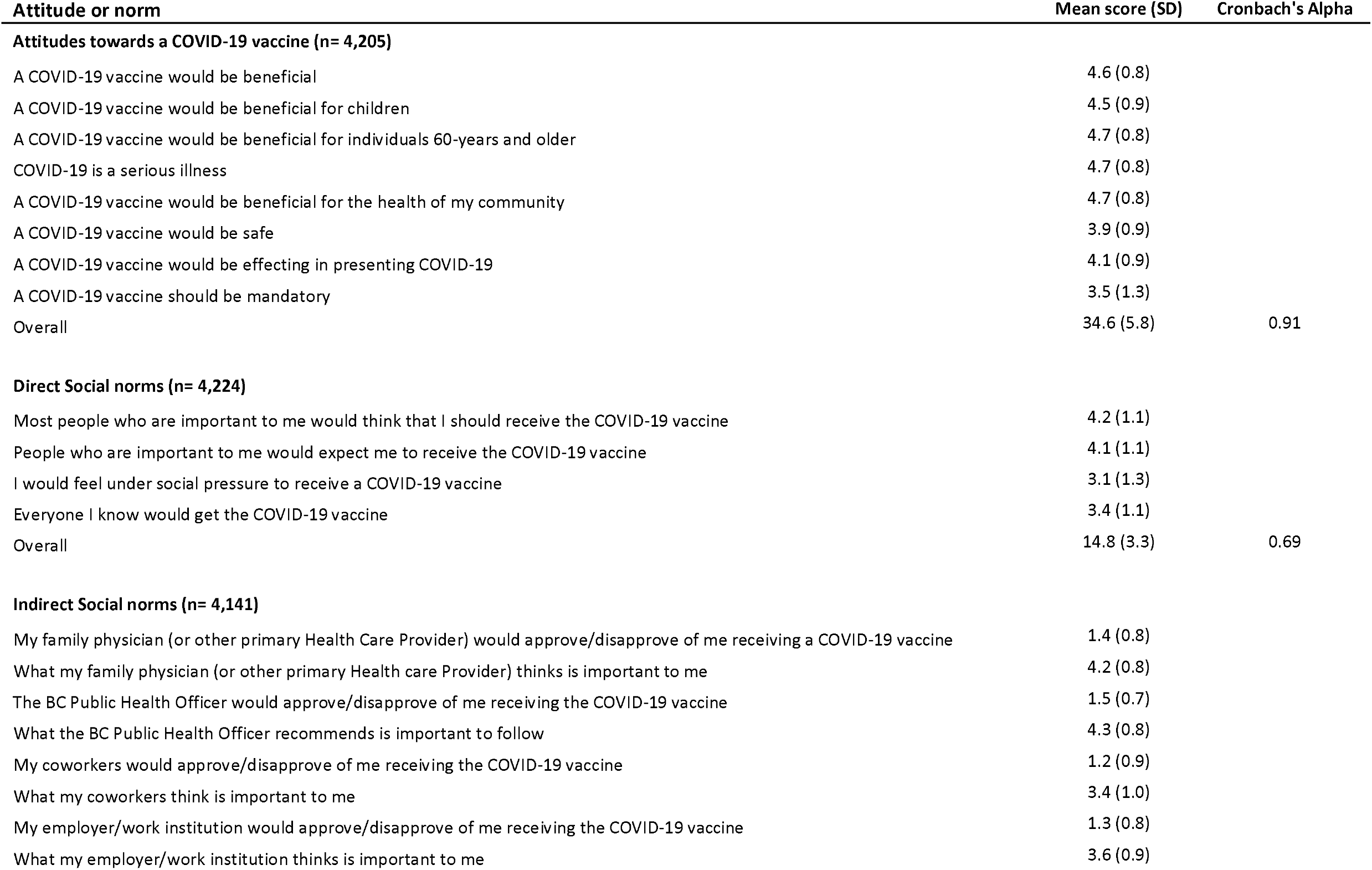
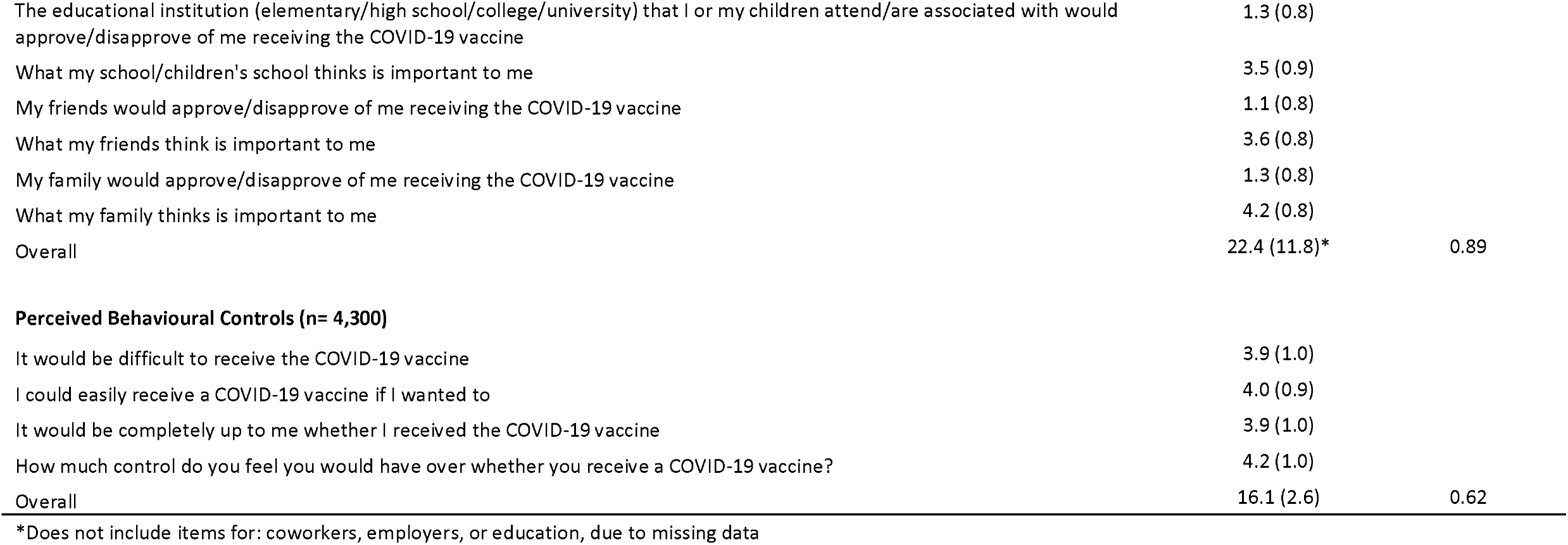
Results of psychological construct scales

**Table 4:**
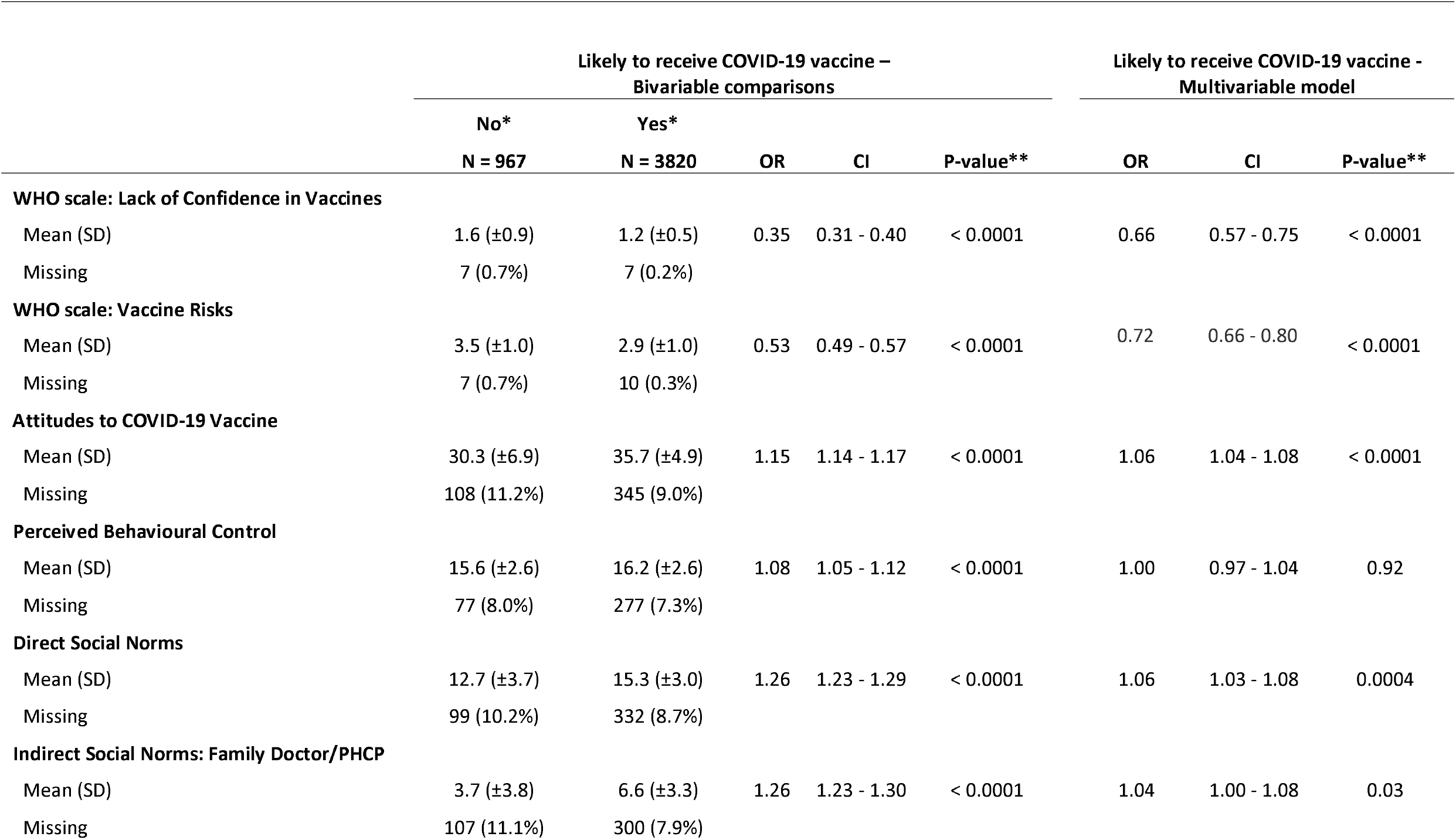
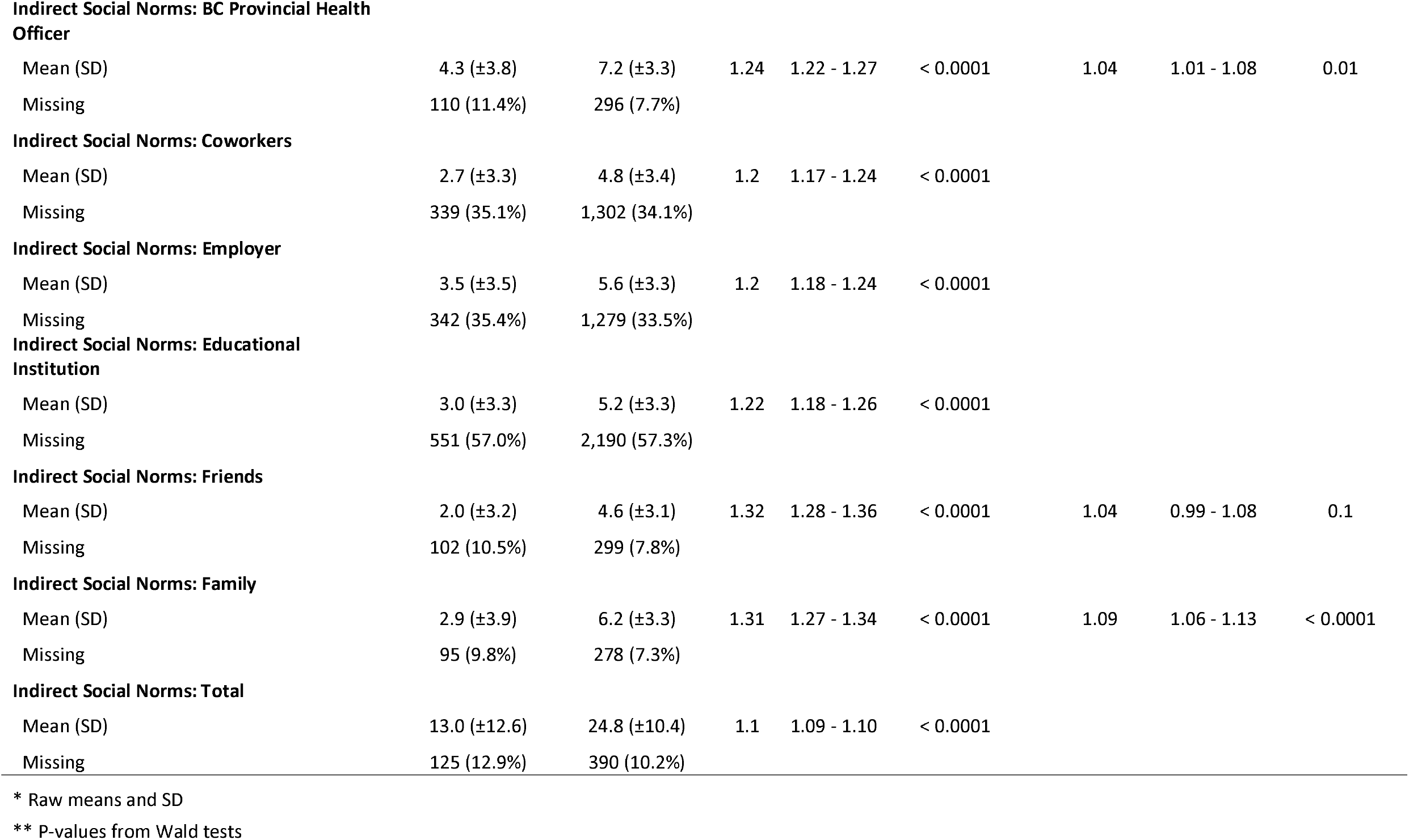
Bivariable comparison and multivariable model of VHS and TPB and intention to receive COVID-19 vaccine (N=4787)

|  | Likely to receive COVID-19 vaccine –<br>Bivariable comparisons |  |  |  |  | Likely to receive COVID-19 vaccine -<br>Multivariable model |  |  |
| --- | --- | --- | --- | --- | --- | --- | --- | --- |
|  | No*<br>N = 967 | Yes*<br>N = 3820 | OR | CI | P-value** | OR | CI | P-value** |
| **WHO scale: Lack of Confidence in Vaccines** |  |  |  |  |  |  |  |  |
| Mean (SD) | 1.6 (±0.9) | 1.2 (±0.5) | 0.35 | 0.31 - 0.40 | < 0.0001 | 0.66 | 0.57 - 0.75 | < 0.0001 |
| Missing | 7 (0.7%) | 7 (0.2%) |  |  |  |  |  |  |
| **WHO scale: Vaccine Risks** |  |  |  |  |  |  |  |  |
| Mean (SD) | 3.5 (±1.0) | 2.9 (±1.0) | 0.53 | 0.49 - 0.57 | < 0.0001 | 0.72 | 0.66 - 0.80 | < 0.0001 |
| Missing | 7 (0.7%) | 10 (0.3%) |  |  |  |  |  |  |
| **Attitudes to COVID-19 Vaccine** |  |  |  |  |  |  |  |  |
| Mean (SD) | 30.3 (±6.9) | 35.7 (±4.9) | 1.15 | 1.14 - 1.17 | < 0.0001 | 1.06 | 1.04 - 1.08 | < 0.0001 |
| Missing | 108 (11.2%) | 345 (9.0%) |  |  |  |  |  |  |
| **Perceived Behavioural Control** |  |  |  |  |  |  |  |  |
| Mean (SD) | 15.6 (±2.6) | 16.2 (±2.6) | 1.08 | 1.05 - 1.12 | < 0.0001 | 1.00 | 0.97 - 1.04 | 0.92 |
| Missing | 77 (8.0%) | 277 (7.3%) |  |  |  |  |  |  |
| **Direct Social Norms** |  |  |  |  |  |  |  |  |
| Mean (SD) | 12.7 (±3.7) | 15.3 (±3.0) | 1.26 | 1.23 - 1.29 | < 0.0001 | 1.06 | 1.03 - 1.08 | 0.0004 |
| Missing | 99 (10.2%) | 332 (8.7%) |  |  |  |  |  |  |
| **Indirect Social Norms: Family Doctor/PHCP** |  |  |  |  |  |  |  |  |
| Mean (SD) | 3.7 (±3.8) | 6.6 (±3.3) | 1.26 | 1.23 - 1.30 | < 0.0001 | 1.04 | 1.00 - 1.08 | 0.03 |
| Missing | 107 (11.1%) | 300 (7.9%) |  |  |  |  |  |  |
| **Indirect Social Norms: BC Provincial Health Officer** |  |  |  |  |  |  |  |  |
| Mean (SD) | 4.3 (±3.8) | 7.2 (±3.3) | 1.24 | 1.22 - 1.27 | < 0.0001 | 1.04 | 1.01 - 1.08 | 0.01 |
| Missing | 110 (11.4%) | 296 (7.7%) |  |  |  |  |  |  |

**Indirect Social Norms: Coworkers**
|  |  |  |  |  |  |
| --- | --- | --- | --- | --- | --- |
| Mean (SD) | 2.7 (±3.3) | 4.8 (±3.4) | 1.2 | 1.17 - 1.24 | < 0.0001 |
| Missing | 339 (35.1%) | 1,302 (34.1%) |  |  |  |

**Indirect Social Norms: Employer**
|  |  |  |  |  |  |
| --- | --- | --- | --- | --- | --- |
| Mean (SD) | 3.5 (±3.5) | 5.6 (±3.3) | 1.2 | 1.18 - 1.24 | < 0.0001 |
| Missing | 342 (35.4%) | 1,279 (33.5%) |  |  |  |

**Indirect Social Norms: Educational Institution**
|  |  |  |  |  |  |
| --- | --- | --- | --- | --- | --- |
| Mean (SD) | 3.0 (±3.3) | 5.2 (±3.3) | 1.22 | 1.18 - 1.26 | < 0.0001 |
| Missing | 551 (57.0%) | 2,190 (57.3%) |  |  |  |
| **Indirect Social Norms: Friends** |  |  |  |  |  |  |  |  |
| Mean (SD) | 2.0 (±3.2) | 4.6 (±3.1) | 1.32 | 1.28 - 1.36 | < 0.0001 | 1.04 | 0.99 - 1.08 | 0.1 |
| Missing | 102 (10.5%) | 299 (7.8%) |  |  |  |  |  |  |
| **Indirect Social Norms: Family** |  |  |  |  |  |  |  |  |
| Mean (SD) | 2.9 (±3.9) | 6.2 (±3.3) | 1.31 | 1.27 - 1.34 | < 0.0001 | 1.09 | 1.06 - 1.13 | < 0.0001 |
| Missing | 95 (9.8%) | 278 (7.3%) |  |  |  |  |  |  |

**Indirect Social Norms: Total**
|  |  |  |  |  |  |
| --- | --- | --- | --- | --- | --- |
| Mean (SD) | 13.0 (±12.6) | 24.8 (±10.4) | 1.1 | 1.09 - 1.10 | < 0.0001 |
| Missing | 125 (12.9%) | 390 (10.2%) |  |  |  |

\* Raw means and SD
\*\* P-values from Wald tests

**Table 5:**
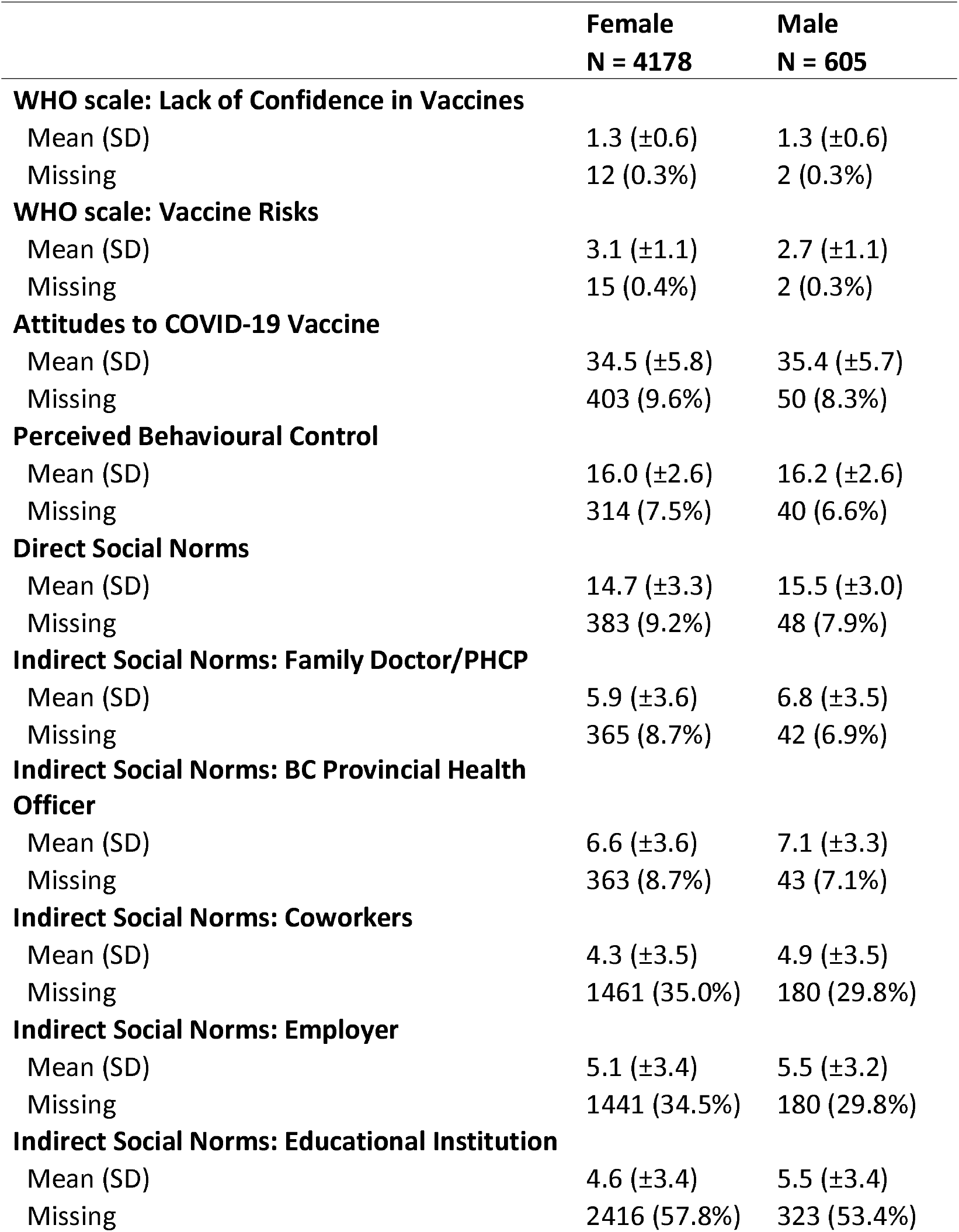
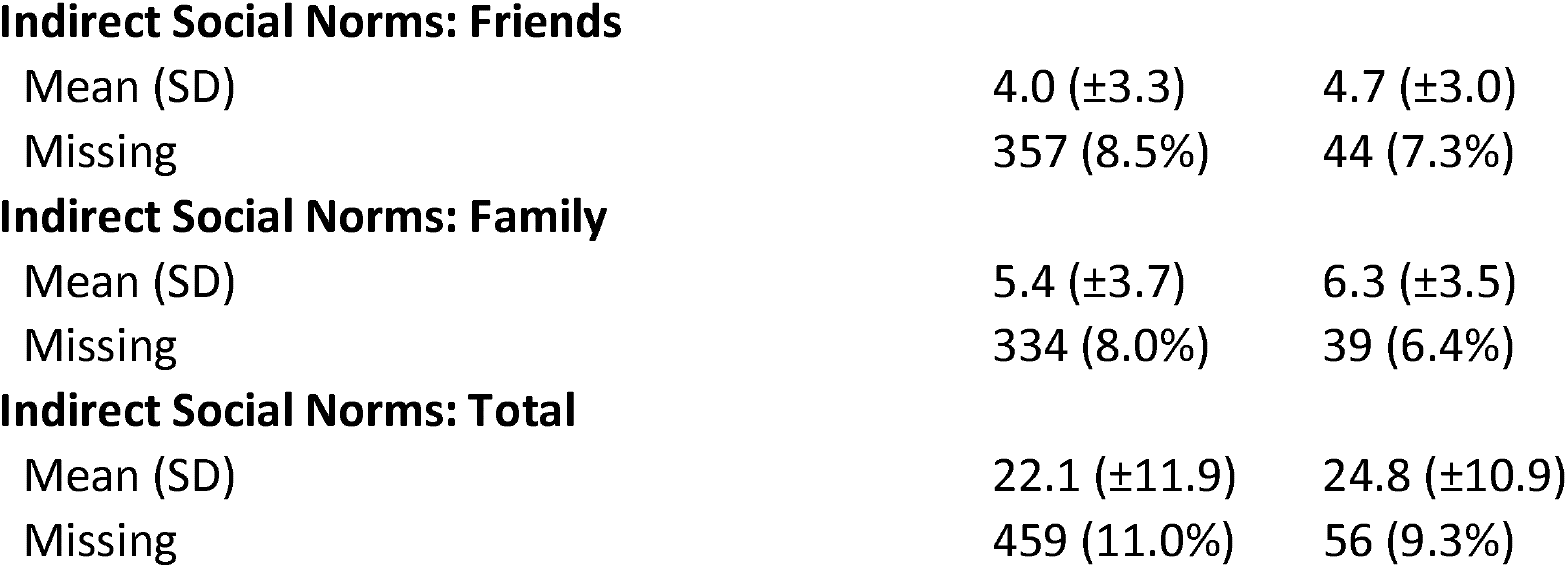
Comparison of psychological constructs (WHO VHS scale and TPB framework) by sex

|  | **Female**<br>**N = 4178** | **Male**<br>**N = 605** |
| --- | --- | --- |
| **WHO scale: Lack of Confidence in Vaccines** |  |  |
| Mean (SD) | 1.3 ($\pm 0.6$) | 1.3 ($\pm 0.6$) |
| Missing | 12 (0.3%) | 2 (0.3%) |
| **WHO scale: Vaccine Risks** |  |  |
| Mean (SD) | 3.1 ($\pm 1.1$) | 2.7 ($\pm 1.1$) |
| Missing | 15 (0.4%) | 2 (0.3%) |
| **Attitudes to COVID-19 Vaccine** |  |  |
| Mean (SD) | 34.5 ($\pm 5.8$) | 35.4 ($\pm 5.7$) |
| Missing | 403 (9.6%) | 50 (8.3%) |
| **Perceived Behavioural Control** |  |  |
| Mean (SD) | 16.0 ($\pm 2.6$) | 16.2 ($\pm 2.6$) |
| Missing | 314 (7.5%) | 40 (6.6%) |
| **Direct Social Norms** |  |  |
| Mean (SD) | 14.7 ($\pm 3.3$) | 15.5 ($\pm 3.0$) |
| Missing | 383 (9.2%) | 48 (7.9%) |
| **Indirect Social Norms: Family Doctor/PHCP** |  |  |
| Mean (SD) | 5.9 ($\pm 3.6$) | 6.8 ($\pm 3.5$) |
| Missing | 365 (8.7%) | 42 (6.9%) |
| **Indirect Social Norms: BC Provincial Health Officer** |  |  |
| Mean (SD) | 6.6 ($\pm 3.6$) | 7.1 ($\pm 3.3$) |
| Missing | 363 (8.7%) | 43 (7.1%) |
| **Indirect Social Norms: Coworkers** |  |  |
| Mean (SD) | 4.3 ($\pm 3.5$) | 4.9 ($\pm 3.5$) |
| Missing | 1461 (35.0%) | 180 (29.8%) |
| **Indirect Social Norms: Employer** |  |  |
| Mean (SD) | 5.1 ($\pm 3.4$) | 5.5 ($\pm 3.2$) |
| Missing | 1441 (34.5%) | 180 (29.8%) |
| **Indirect Social Norms: Educational Institution** |  |  |
| Mean (SD) | 4.6 ($\pm 3.4$) | 5.5 ($\pm 3.4$) |
| Missing | 2416 (57.8%) | 323 (53.4%) |
| **Indirect Social Norms: Friends** |  |  |
| Mean (SD) | 4.0 ($\pm 3.3$) | 4.7 ($\pm 3.0$) |
| Missing | 357 (8.5%) | 44 (7.3%) |
| **Indirect Social Norms: Family** |  |  |
| Mean (SD) | 5.4 ($\pm 3.7$) | 6.3 ($\pm 3.5$) |
| Missing | 334 (8.0%) | 39 (6.4%) |
| **Indirect Social Norms: Total** |  |  |
| Mean (SD) | 22.1 ($\pm 11.9$) | 24.8 ($\pm 10.9$) |
| Missing | 459 (11.0%) | 56 (9.3%) |

## DISCUSSION

To support COVID-19 vaccine implementation, this study investigated intention to receive the COVID-19 vaccine and determine predictors of COVID-19 vaccine uptake of adults living in BC. The majority (79.8%) of adults surveyed intend to receive a COVID-19 vaccine if available to the public and recommended for them. In multivariable modeling, older individuals (>60 years) were more likely to intend to receive the COVID-19 vaccine. However, other key populations including essential non-health care workers, those who identified as non-white or Indigenous, as well as those with less than high school education indicated that they are less likely to intend to receive a COVID-19 vaccine. From the WHO and TPB scales, we found that those who report higher lack of confidence in vaccines and higher perceived risk of vaccines were less likely to indicate an intention to vaccinate. As well, overall attitudes to vaccines and social norms were significant predictors of intention to receive a COVID-19 vaccine. When we adjusted the multivariable models for sex, only negligible changes were observed in the results (data not shown). Understanding population-level intention to receive the COVID-19 vaccine is critical to success of COVID-19 vaccination programs and ultimately COVID-19 pandemic control.^2^

Our study findings are consistent with results from unpublished polls in Canada investigating COVID-19 vaccine intention, which have found that the majority of Canadians intend to get a COVID-19 vaccine when it becomes available.^2,27,28^ In addition, our findings across demographic subgroups align with published trends in the United States, reporting that women, those with a high school education or less, younger adults (<65 years), and those who identify as Black are less likely to report intention to receive a COVID-19 vaccine.^29^ A study published in the United Kingdom reported similar findings by age: older age was significantly associated with increased likelihood of vaccination.^30^ However, our findings contrast overall vaccine intention polls from the United States and the UK, which indicate that 57.6% and 53% of the population in those countries respectively intended to be vaccinated against COVID-19^31,32^

Results from the WHO VHS and the TPB scales provide important insights to guide public health programming (Table 4). Specifically, to instill confidence in COVID-19 vaccines, messaging should focus on the benefit of vaccines, including their impact on society overall and benefits to children, community, families and individuals. Information on vaccine safety from vaccine trials, as well as plans for ongoing, transparent monitoring and reporting of side effects of COVID-19 vaccines should be broadly shared to strengthen public confidence. As our findings show that family physicians and Provincial Health Officers are influential in vaccine decision making, public health leadership needs to utilize their influence to optimize COVID-19 vaccine coverage.

Leading health authorities, such as the WHO, have identified the need to ensure that everyone is protected by full immunization regardless of socioeconomic and gender-related barriers.^33^ While findings from our study indicate older individuals are more likely to intend to receive the vaccine, in the priority populations identified by ACIP and NACI, the intention to vaccinate is not near to 100%. Priority populations, including essential non-health care workers, non-white, and Indigenous populations are less likely to intend to receive the vaccine, indicating that strong vaccine messaging is still needed. Public health leaders need to work specifically with these communities, to better understand their concerns and build confidence and trust in the vaccine program.

Additional Canada-wide research is needed to understand vaccine intentions, to determine if vaccine intentions vary by province. Future research investigating COVID-19 vaccine intentions should continue to incorporate validated scales and established theoretical frameworks like the VHS and TPB, rather than relying on unvalidated online polls.

### Limitations

This study included a population of individuals who were recruited from large health research cohorts, and has a higher percentage of respondents who identified as female, white, with more than high school education, and were more likely to live in the southern part of the province compared to the general population of BC.^34^ While we had a lower than expected response rate, there was no observed differences in age distribution between the responders and non-responders.

### Conclusion

Our study shows that while the majority of respondents intend to receive the COVID-19 vaccine, there are important factors associated with intention to vaccinate, which can guide vaccination policies and immunization programs, and provide valuable recommendations for vaccine priority groups captured in our study sample, specifically older age groups, health care workers and other essential workers. To our knowledge, this is the first study that has drawn from a large provincial sample in Canada, using established and rigorous theoretical frameworks to investigate intention to receive a COVID-19 vaccine.

## Data Availability

The datasets generated and analyzed during the current study are not publicly available. At the time of consent participants were not advised that data might be made publicly available, and so we felt it was not appropriate to make the data publicly available. Aggregate data may be made available by the corresponding author upon reasonable request.

## Declarations

### Ethics approval and consent to participate

Ethical approval was received from The University of British Columbia Research Ethics Board (H20-01421). All methods performed as a part of this study were in accordance with the UBC Research Ethics Board guidelines. Informed consent to participate was obtained from participants.

### Consent for publication

Not applicable.

### Availability of data and materials

The datasets generated and analysed during the current study are not publicly available. At the time of consent participants were not advised that data might be made publicly available, and so we felt it was not appropriate to make the data publicly available. Aggregate data may be made available by the corresponding author upon reasonable request.

## Acknowledgements

Not applicable.

