## Supplemental Material 1 for "Intention to receive a COVID-19 vaccine: Results from a population-based survey in Canada"

Additional File 1:

Survey Questions

Core Module

1. Are you 25-69 years old (inclusive)?
   1. Yes
   2. No (End Survey)
2. Are you a resident of BC?
   1. Yes
   2. No (End survey)

The next series of questions ask about presence or absence of symptoms for COVID-19. Symptoms of COVID-19 may include

1) Fever >= 38
2) Feverishness
3) Chills
4) Cough
5) Fatigue
6) Muscle pain
7) Joint pain
8) Sore throat
9) Nasal congestion
10) Shortness of breath
11) Wheezing
12) Other respiratory symptom
13) Headache
14) Alteration to taste
15) Alteration to smell
16) Nausea/vomiting
17) Abdominal pain
18) Diarrhea.
3. Do you think you have had COVID-19?

- No
- Yes

4. *If yes to question #3,* Why do you think you have had COVID-19? Please check all that apply

- Symptoms (cough, fever, shortness of breath, etc.)
- Nasal/throat test result
- Health care provider
- Contact with case

5. *If yes to question #3,* Where do you believe you came into contact with a case of COVID-19? If you are unsure where, please choose the most likely exposure environment.

- Household
- High risk environment (including healthcare setting)
- Occupational setting
- Other local setting – Specify:
- Travel
- Don’t know

6. *If yes to question #3,* Did you have a nasal/throat swab to test for COVID-19?

- Yes
- No

7. *If yes to question #6,* What month did you have the COVID-19 test?
   - January
   - February
   - March
   - April
   - May
   - June
   - July
   - August
   - September
   - October
   - November
   - December
   - January 2021
8. *If yes to question #6,* Was it early, mid, or late [**month from above**]?

- Early
- Mid
- Late

9. *If yes to question #6,* What was the result of your COVID-test?

- Positive
- Negative
- I have not received the results yet

10. *If “positive” to question #9,* Did you require hospitalization due to COVID-19?

- No
- Yes

11. *If yes to question #10,* What was your admission date? *If you are not sure, leave blank*
12. *If yes to question #10,* What was your discharge date? *If you are not sure, leave blank*
13. *If yes to question #10*, Did you require ventilation because of COVID-19?

- Yes
- No

14. *If yes to question #3,* What were your symptoms while you were sick/thought you were sick? *Select all that apply*

- Fever
- Cough
- Fatigue
- Muscle pain
- Sore throat
- Shortness of breath
- Headache
- Decreased sense of smell
- Nausea/vomiting
- Diarrhea
- Other symptoms

15. *If endorses any in question #14,* Approximately when did the symptoms start?

- January
- February
- March
- April
- May
- June
- July
- August
- September
- October
- November
- December
- January 2021

16. *If endorses any in question #14,* Approximately when during [**pipe in from above**] did the symptoms begin?

- Early
- Mid
- Late

17. *If yes to question #3,* Did you require any over the counter medication for any symptoms (cough, fatigue, fever, pain)?

- Yes
- No

18. *If endorsed any in question #14,* Are your symptoms resolved?

- Yes
- No

19. *If yes to question #3,* Are you currently in isolation?

- Yes
- No

20. *If yes to question #3,* Where did you stay during the time you were sick/thought you were sick? *Check all that apply*

- I did not change where I was staying
- In home isolation (separated from other household members)
- In home isolation (not separated)
- In a rental or hotel
- In a hospital
- Other

21. *If “contact with case” in question #4*, Where did your contact with a known COVID-19 positive person occur?

- Household
- High risk environment (including a health care setting)
- Occupational setting
- Other local setting
- Travel

22. *If “high risk environment” in question #21*, In what high risk environment do you believe you came in contact with a COVID-19 positive person?

- Hospital
- Nursing home
- Public health clinic
- Shelter
- COVID testing facility
- Other – specify:

23. *If “occupational setting” in question #21*, What occupational risk may have caused you to be in contact with a COVID-19 positive individual?

- health care worker - direct patient contact,
- health care worker - indirect patient contact,
- law enforcement or first responder,
- vulnerable population service provider,
- food and agricultural service provider (grocery, pharmacy, liquor,

other household products),

- transportation worker (bus driver, sky train operators, west coast express, seabus, handi-dart, customer facing airport employees, etc)
- Childcare worker
- Correctional officer
- Teacher/other school staff
- Hairdresser/Barber
- Aesthetician
- Flight attendant
- Factory worker

24. Have you traveled outside of British Columbia since January 2020?

- Yes
- No

25. *If yes to question #24,* Where and how many times you have traveled since January 2020? Check all that apply.

- I traveled outside of BC (within Canada) once
- I traveled outside of BC (within Canada) multiple times
- I traveled outside of Canada once
- I traveled outside of Canada multiple times

26. What is your current age (years)?
27. What sex were you assigned at birth?

- Female
- Male
- Intersex
- Prefer to self-describe – Specify:
- Prefer not to answer

28. Which best describes your current gender identity?

- Man
- Woman
- Two-Spirit
- Non-Binary, GenderQueer, Agender, or similar identity
- Prefer to self-describe – Specify:
- Prefer not to answer

29. *If question #27 and question #28 are not [“male” and “man”] or [“female” and “woman”],* What gender do you currently live as in your day-to-day life?

- Man
- Woman
- Sometimes man, sometimes woman
- Something other than man or woman

30. What is your sexual orientation?

- Asexual
- Bisexual
- Demisexual
- Gay/Lesbian
- Heterosexual
- Pansexual
- Prefer to self-describe – specify: ______
- Prefer not to answer

31. Are you an Indigenous person originating from North America? *Check any that apply.*

- First Nation
- Métis
- Inuit
- Non-status First Nations
- Other Indigenous
- Prefer not to answer
- I am not an Indigenous person originating from North America

32. *If endorse any Indigenous group in question #31,* Do you live on or off reserve?

- On-reserve
- Off-reserve
- Prefer not to answer

33. Do you consider yourself to be? *Please check all that apply to you.*

- Black African (e.g., Nigerian, Somali)
- Black Caribbean (e.g., Haitian)
- Black Other (e.g., Black Canadian)
- White
- Chinese or Taiwanese
- Filipino
- Japanese
- Korean
- Latin American (e.g., Chilean, Costa Rican, Mexican)
- South Asian (e.g., Indian, Bangladeshi, Pakistani, Punjabi, and Sri Lankan)
- Southeast Asian (e.g.,Cambodian, Laotian, Malaysian, Vietnamese)
- Arab (e.g., Egyptian, Kuwaiti, and Libyan)
- West Asian (e.g. Iraqi, Isreali, Lebanese, Afghani, Iranian)
- Central Asian (e.g., Kazakhstani, Krgyzstani, Tajikistani, Turkmenistani)
- Multiple races / Multiracial / “Mixed”
- Prefer not to answer
- Other, please specify: _____________________________

34. What is your current legal status in Canada?

- Canadian citizen – born in Canada
- Naturalized Canadian citizen
- Landed immigrant/Permanent Resident
- Refugee/Protect Person/Refugee claimant
- Here with Temporary Work Papers
- Here with Humanitarian and Compassionate approval
- Here as a visitor
- Here on a Student Visa
- Undocumented/Illegal Immigrant
- Other – Please specify:
- Don't know
- Prefer not to answer

35. *If “landed immigrant” in question #34,* in what year did you first become a landed immigrant to Canada? Please enter the year as 4 digits (YYYY).

- YYYY

36. What is your marital status?

- Never legally married – currently single
- Never legally married – living with partner < 12 months
- Never legally married – in a relationship
- Legally married
- Separated, but still legally married
- Common law
- Divorced
- Widowed

37. How many adults (18+ years) live in your household?
38. How many children under the age of five live in your household?
39. How many children age five to seventeen live in your household?
40. What is your highest achieved education level?

- No certificate, diploma or degree
- Secondary (high) school diploma or equivalency certificate
- Apprenticeship or trades certificate or diploma
- College, CEGEP or other non-university certificate or diploma
- University certificate or diploma below bachelor level
- University certificate, diploma or degree at bachelor level or above

41. Are you currently attending school, college, university or other?

- Yes
- No

42. Do you expect to be a student this fall?

- Yes
- No

43. *If “yes” to question #42,* Do you expect to be a full-time or part-time student in the fall?

- Full-time
- Part-time
- Don’t know

44. Do you consider yourself an essential worker? *Will direct to either question #45 or #46*

- No
- Yes, health worker
- Yes, other essential worker

45. *If “health worker” in question #44,* Type of health worker:

- Physician
- Nurse
- Occupational Therapist
- Physical Therapist
- Respiratory Therapist
- Care Aid/Personal Care Worker
- Dietician
- Healthcare worker - indirect patient contact (stocking shelves, cleaners, cafeteria, mechanics/engineers, etc)
- other – specify:

46. *If “Yes, other essential worker” in question #44,* Type of other essential worker:

- Law enforcement or first-responder
- Vulnerable population service provider (e.g. social worker, youth worker)
- Food and agriculture service provider (grocery, pharmacy, liquor, other household products)
- Transportation worker (bus driver, sky train operations, west coast express, seabus, handi-dart, customer facing airport employees, etc)
- Engineer or construction worker
- other – specify

47. The section asks about whether you've ever been diagnosed with a chronic health condition. We are interested in long-term conditions which are expected to last or have already lasted 6 months or more and that have been diagnosed by a health professional. Please check all that apply.

    Has a doctor ever told you that you have...

|  | Yes | No | Prefer not to answer |
| --- | --- | --- | --- |
| Asthma (A common lung disorder in which inflammation causes the bronchi to swell and narrow the airways, creating breathing difficulties) |  |  |  |
| COPD or emphysema (Chronic Obstructive Pulmonary Disease or COPD a long-term, progressive disease of the lungs that primarily causes shortness of breath due to over-inflation of the alveoli (air sacs in the lung)) |  |  |  |
| Chronic lung disease (A disorder that can be caused by smoking tobacco or inhaling air pollutants that usually develops slowly, may get worse over time, and affects the lungs and other parts of the respiratory system) |  |  |  |
| Insulin resistance/pre-diabetes/borderline diabetes (A condition in which a decreased response to insulin by a person's body tissues results in the production of larger quantities of insulin to maintain normal levels of glucose in the blood) |  |  |  |
| Diabetes (A disease in which there is reduced production of, or response to, insulin so that glucose cannot be absorbed into the cells of the body) |  |  |  |
| High blood pressure/hypertension (Abnormally high arterial blood pressure that is usually indicated by an adult systolic blood pressure of 140 mm Hg or greater or a diastolic blood pressure of 90 mm Hg or greater) |  |  |  |
| Heart disease (A disease in which an accumulation of plaque in the vessels that supply blood to the heart causes impaired heart functioning) |  |  |  |
| Coronary Artery Disease/Myocardial Infarction/Heart Attack (Deprivation of oxygen, usually due to blockage of a diseased coronary artery, damages the heart and causes chest pain) |  |  |  |
| Heart Failure (A condition in which the heart fails to pump adequate amounts of blood to body tissues, resulting in an accumulation of blood returning to the heart from the veins) |  |  |  |
| Cardiac arrhythmia/Atrial Fibrillation (A condition in which the normal rhythmical contractions of the heart are replaced by rapid irregular twitchings of the muscular wall) |  |  |  |
| Stroke (Sudden impairment or loss of consciousness, sensation, and voluntary motion that is caused by rupture or obstruction of a blood vessel supplying the brain and is accompanied by permanent damage of brain tissue) |  |  |  |
| Deep vein thrombosis/Pulmonary Embolism (A condition marked by the formation of a clot within a deep vein that may be accompanied by symptoms and that is potentially life threatening if the clot dislodges and obstructs the pulmonary vasculature) |  |  |  |
| Peripheral Vascular Disease (A disease affecting blood vessels outside of the heart and especially those vessels supplying the extremities) |  |  |  |
| High cholesterol (Too much cholesterol in the blood, usually considered above 6.2 mmol/L) |  |  |  |
| Liver disease (i.e., fatty liver) (An abnormal condition of the liver that is marked by excess fat accumulation in the liver cells and that is typically associated with obesity, malnutrition, rapid weight loss, excessive alcohol consumption, high blood fat content, or diabetes) |  |  |  |
| Liver cirrhosis (Compromised liver structure and function that results from liver cell death, inflammation, and scar tissue) |  |  |  |
| Renal problem/Kidney problem. (Impaired ability of the kidneys to perform vital functions that can be due to an acute event or a chronic condition or disease) |  |  |  |
| Autoimmune Disorder (A disorder caused by a reaction of the immune system against the organs or tissues of the body) |  |  |  |
| Pneumonia that was confirmed using chest x-rays (An acute disease that is marked by inflammation of the lung tissue and typically causes fever, chills, cough, difficulty breathing, fatigue, chest pain, and reduced lung expansion) |  |  |  |
| Chronic neurological or neuromuscular disorder (A long-lasting disorder that is caused by a dysfunction in part of the brain or nervous system and that results in physical or psychological symptoms or that affects voluntary muscle control) |  |  |  |

48. Are you a person living with a disability?

- Yes
- No
- Prefer not to answer

49. Are you a person living with HIV?

- Yes
- No
- Don’t know
- Prefer not to answer

50. Has a doctor ever told you that you have cancer (excluding skin malignancies)?

- Yes
- No
- Prefer not to answer

51. *If yes to question #50*, What type(s) of cancer were you diagnosed with? Please select all that apply.

- Anal
- High Grade Anal precancer (Anal Intraepithelial Neoplasia, AIN 2 or 3)
- Bladder
- Bone
- Breast
- Cervical (“HPV”)
- High Grade Cervical precancer * (Cervical Intraepithelial Neoplasia or CIN 2 OR 3)
- Colon or Rectum
- Endometrial (ie. of the uterus)
- Kaposi Sarcoma
- Kidney
- Liver
- Lung
- Lymphoma / leukemia
- Oral or pharynx
- Ovarian
- Skin (melanoma, basal, squamous cells)
- Stomach or Small Bowel
- Thyroid
- Vulvar
- High Grade Vulvar or vaginal precancer (Vulvar or Vaginal Intaepithelial Neoplasia, VIN or VaIN 2 or 3)
- Other, please specify:
- Don't know
- Prefer not to answer

52. During the COVID-19 pandemic (mid March to now) were you due for, or in need of, any screening appointments? *Select all that apply.*

- Pap test
- Colonoscopy
- Mammogram
- Other, please specify: ______

53. *If any checked off in question #52,* Did you attend the above appointments for which you were due?

- I attended all
- I attended some
- I attended none
- Prefer not to answer

54. *If some/none,* Why did you not attend the appointments?

- My doctor or clinic was not accepting in-person appointments
- My appointment was considered non-urgent
- Worried about visiting doctor or doctors’ office
- Other, please specify: _____________

55. Are you pregnant?

- Yes
- No

56. *If yes to above*, How many weeks pregnant are you? *If you don’t know, please leave blank.*
57. *If no to question #55,* Have you been pregnant at anytime since March 2020?

- Yes
- No

58. *If yes to question #57,* Are you 6 weeks or less postpartum?

- Yes
- No

53. *If yes to question #57,* Since March 2020 have you suffered from any miscarriages?

- Yes
- No

54. Please enter your height in feet and inches
55. Please enter your weight in pounds

We would now like to ask you some questions about your feelings surrounding vaccines, and whether or not your thoughts about vaccines for your health have changed since the COVID-19 Pandemic

56. If a COVID-19 vaccine were to become available to the public, and recommended for you, how likely are you to receive it?

- Very unlikely
- Unlikely
- Neutral
- Somewhat likely
- Very likely

57. Do you receive all recommended vaccines when offered to you by health care professionals?

- Yes
- No
- Don’t know

58. Thinking back to before the beginning of the pandemic (December 2019), how much did you value vaccines?

- No value
- Very little value
- Neutral
- A little value
- Valued a lot

59. Today, how much do you value vaccines?

- No value
- Very little value
- Neutral
- A little value
- Valued a lot

60. Have you received the influenza vaccine in the past 5 years?

- Never
- 1-2 times
- 3-4 times
- Every year
- Don’t know

61. Do you intend to receive the influenza vaccine in the fall of 2020?

- Yes
- No
- Don’t know
- I have already received the vaccine

62. *WHO Vaccine Hesitancy Scale (Vaccine Confidence Scale)* [Section Instructions:] *This section will ask you about your opinion about vaccines, in general.* *Please indicate how much you agree or disagree with each statement below.*

|  | Strongly disagree | Somewhat disagree | Neutral | Somewhat agree | Strongly agree |
| --- | --- | --- | --- | --- | --- |
| Childhood vaccines are important for a child’s health |  |  |  |  |  |
| Getting vaccines is a good way to protect children from disease |  |  |  |  |  |
| Childhood vaccines are effective |  |  |  |  |  |
| Having a child vaccinated is important for the health of others in my community |  |  |  |  |  |
| All childhood vaccines offered by the BC immunization program in my community are beneficial |  |  |  |  |  |
| The information I receive about vaccines from the vaccination program is reliable and trustworthy |  |  |  |  |  |
| Generally, I do what my doctor or health care provider recommends about vaccines |  |  |  |  |  |
| New vaccines carry more risks than older vaccines |  |  |  |  |  |
| I am concerned about potential serious adverse effects of vaccines |  |  |  |  |  |
| Children do not need vaccines for diseases that are not common anymore |  |  |  |  |  |
| The risk associated with diseases is larger than the risk associated with vaccinations |  |  |  |  |  |

Vaccine Module

*Assuming that a safe and effective COVID-19 vaccine was available and recommended, please indicate how much you agree or disagree with the following statements:*

Attitudes

[*5-point scale: 1 strongly disagree, 3 neutral, 5 strongly agree]*

63. A COVID-19 vaccine would be beneficial
64. A COVID-19 vaccine would be beneficial for children
65. A COVID-19 vaccine would be beneficial for individuals 60-years and older
66. COVID-19 is a serious illness
67. A COVID-19 vaccine would be beneficial for the health of my community
68. A COVID-19 vaccine would be safe
69. A COVID-19 vaccine would be effective in preventing COVID-19
70. A COVID-19 vaccine should be mandatory

*Please answer the following questions, with the assumption that a safe and effective COVID-19 vaccine is available and recommended.*

Social Norms

Q77-80, 82, 84, 86, 88, 90, 92, 94: [*5-point scale: 1 strongly disagree, 3 neutral, 5 strongly agree]*

Q81, 83, 85, 87, 89, 91, 93: [*5-point scale: 1 strongly disapprove, 3 neutral, 5 strongly approve]*

71. Most people who are important to me would think that I should receive the COVID-19 vaccine
72. People who are important to me would expect me to receive the COVID-19 vaccine
73. I would feel under social pressure to receive a COVID-19 vaccine
74. Everyone I know would get the COVID-19 vaccine
75. My family physician (or other primary Health Care Provider) would approve/disapprove of me receiving a COVID-19 vaccine
76. What my family physician (or other primary Health Care Provider) thinks is important to me
77. The BC Public Health Officer would approve/disapprove of me receiving the COVID-19 vaccine
78. What the BC Public Health Officer recommends is important to follow
79. My coworkers would approve/disapprove of me receiving the COVID-19 vaccine
80. What my coworkers think is important to me
81. My employer/work institution would approve/disapprove of me receiving the COVID-19 vaccine
82. What my employer/work institution thinks is important to me
83. The educational institution (elementary/high school/college/university) that I or my children attend/are associated with would approve/disapprove of me receiving the COVID-19 vaccine
84. What my school/children’s school thinks is important to me
85. My friends would approve/disapprove of me receiving the COVID-19 vaccine
86. What my friends think is important to me
87. My family would approve/disapprove of me receiving the COVID-19 vaccine
88. What my family thinks is important to me

Behavioural Control

Q95-98: [*5-point scale: 1 strongly disagree, 3 neutral, 5 strongly agree]*

Q99: [*Sliding scale: 1 numerous, 5 some, 10 very few]*

For the next 3 questions: If a COVID-19 vaccine was offered and publicly funded and available, like the flu shot …

89. … It would be difficult to receive the COVID-19 vaccine
90. … I could easily receive a COVID-19 vaccine if I wanted to
91. … It would be completely up to me whether I received the COVID-19 vaccine
92. How much control do you feel you would have over whether you receive a COVID-19 vaccine?

- Very little control
- Not much control
- Neutral
- Some control
- A lot of control

93.
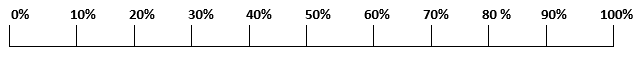
The number of events outside my control which would prevent me from having a COVID-19 vaccination are:

**0	5	10**
