## Supplemental Material 2 for "Intention to receive a COVID-19 vaccine: Results from a population-based survey in Canada"

Additional File 2:

Scale Items

VHS Sub-scale 1: Lack of Confidence

1. Childhood vaccines are important for a child’s health
2. Getting vaccines is a good way to protect children from disease
3. Childhood vaccines are effective
4. Having a child vaccinated is important for the health of others in my community
5. All childhood vaccines offered by the BC immunization program in my community are beneficial
6. The information I receive about vaccines from the vaccination program is reliable and trustworthy
7. Generally, I do what my doctor or health care provider recommends about vaccines

VHS Sub-scale 2: Risks

1. New vaccines carry more risks than older vaccines
2. I am concerned about potential serious adverse effects of vaccines

TPB: Attitudes

1. A COVID-19 vaccine would be beneficial
2. A COVID-19 vaccine would be beneficial for children
3. A COVID-19 vaccine would be beneficial for individuals 60-years and older
4. COVID-19 is a serious illness
5. A COVID-19 vaccine would be beneficial for the health of my community
6. A COVID-19 vaccine would be safe
7. A COVID-19 vaccine would be effective in preventing COVID-19
8. A COVID-19 vaccine should be mandatory

TPB: Direct Social Norms

1. Most people who are important to me would think that I should receive the COVID-19 vaccine
2. People who are important to me would expect me to receive the COVID-19 vaccine
3. I would feel under social pressure to receive a COVID-19 vaccine
4. Everyone I know would get the COVID-19 vaccine

TPB: Indirect Social Norms

1. My family physician (or other primary Health Care Provider) would approve/disapprove of me receiving a COVID-19 vaccine
2. What my family physician (or other primary Health Care Provider) thinks is important to me
3. The BC Public Health Officer would approve/disapprove of me receiving the COVID-19 vaccine
4. What the BC Public Health Officer recommends is important to follow
5. My coworkers would approve/disapprove of me receiving the COVID-19 vaccine
6. What my coworkers think is important to me
7. My employer/work institution would approve/disapprove of me receiving the COVID-19 vaccine
8. What my employer/work institution thinks is important to me
9. The educational institution (elementary/high school/college/university) that I or my children attend/are associated with would approve/disapprove of me receiving the COVID-19 vaccine
10. What my school/children’s school thinks is important to me
11. My friends would approve/disapprove of me receiving the COVID-19 vaccine
12. What my friends think is important to me
13. My family would approve/disapprove of me receiving the COVID-19 vaccine
14. What my family thinks is important to me

TPB: Perceived Behavioural Controls

For the next 3 questions: If a COVID-19 vaccine was offered and publicly funded and available, like the flu shot …

1. … It would be difficult to receive the COVID-19 vaccine
2. … I could easily receive a COVID-19 vaccine if I wanted to
3. … It would be completely up to me whether I received the COVID-19 vaccine

4) How much control do you feel you would have over whether you receive a COVID-19 vaccine?
