## Supplemental Material 3 for "Intention to receive a COVID-19 vaccine: Results from a population-based survey in Canada"

APPENDIX C:

Reporting Guideline Checklist

Burns, KEA, Kho ME. How to assess a survey report: a guide for readers and peer reviewers.

CMAJ [Internet]. 2015 Apr [cited 2020 Oct 30] 187(6): E198-E205. Available from:[10.1503/cmaj.140545](https://dx.doi.org/10.1503%2Fcmaj.140545)

**A guide for appraising survey reports**

1. Was a clear research question posed? **Lines 92-97.**
   - 1a. Does the research question or objective specify clearly the type of respondents, the topic of interest, and the primary and secondary research questions to be addressed?  **Lines 92-97.**
2. Was the target population defined **Line 94-95**, and was the sample representative of the population? **Lines 107-108, 156-157.**
   - 2a. Was the population of interest specified?  **Line 94-95.**
   - 2b. Was the sampling frame specified? **Lines 107-108.**
3. Was a systematic approach used to develop the questionnaire? **Lines 128-138.**
   - 3a. *Item generation and reduction:* Did the authors report how items were generated and ultimately reduced? **Lines 128-130.**
   - 3b. *Questionnaire formatting:* Did the authors specify how questionnaires were formatted? **135-136, 161-165, and Supplemental Material 1.**
   - 3c. *Pretesting:* Were individual questions within the questionnaire pretested?  **Lines 136-138.**
4. Was the questionnaire tested? **Lines 136-138.**
   - 4a. *Pilot testing:* Was the entire questionnaire pilot tested?  **Line 137.**
   - 4b. *Clinimetric testing:* Were any clinimetric properties (face validity or clinical sensibility testing, content validity, inter- or intra-rater reliability) evaluated and reported?  **Line 137.**
5. Were questionnaires administered in a manner that limited both response and nonresponse bias? **Lines 110-114.**
   - 5a. Was the method of questionnaire administration appropriate for the research objective or question posed? **Lines 102-104.**
   - 5b. Were additional details regarding prenotification **(n/a)**, use of a cover letter **(lines 109)** and an incentive for questionnaire completion provided? **(lines 114).**
6. Was the response rate reported **(lines 186-189)**, and were strategies used to optimize the response rate **(lines 110-114)**?
   - 6a. Was the response rate reported (alternatively, were techniques used to assess nonresponse bias)?  **Lines 186-189.**
   - 6b. Was the response rate defined?  **Lines 146-150.**
   - 6c. Were strategies used to enhance the response rate (including sending of reminders)?  **Lines 110-114.**
   - 6d. Was the sample size justified?  **Lines 155-156.**
7. Were the results clearly and transparently reported? **Lines 184-239, Tables 2-5.**
   - 7a. Does the survey report address the research question(s) posed or the survey objectives?  **Lines 204-205.**
   - 7b. Were methods for handling missing data reported?  **Line 177-179.**
   - 7c. Were demographic data of the survey respondents provided?  **Lines 195-202, and Table 1.**
   - 7d. Were the analytical methods clear?  **Lines 159-181.**
   - 7e. Were the results succinctly summarized?  **Lines 184-239.**
   - 7f. Did the authors’ interpretation of the results align with the data presented?  **Lines 195-239, and Tables 2-5.**
   - 7g. Were the implications of the results stated? **Lines 267-285.**
   - 7h. Was the questionnaire provided in its entirety (as an electronic appendix or in print)?  **Supplemental Material 1.**
